# Testing the effects of the timing of application of preventative procedures against COVID-19: An insight for future measures such as local emergency brakes

**DOI:** 10.1101/2020.06.02.20120352

**Authors:** Francis Scullion, Geraldine Scullion

## Abstract

As many countries plan to lift lockdown measures aimed at suppression of COVID-19, data from early regional epidemics in Italy were analysed to ascertain the effectiveness of the timing of preventative measures. The cumulative caseload data were extracted from regional epidemics in Italy. Epidemic features in regions where lockdown was applied early were compared to those where lockdown was applied later for statistical differences. There were statistically significant differences in the timing of the peak, the cumulative incidence at peak and the case/km^2^ at peak between regions where the lockdown had been applied early and those where it was applied late. The peak occurred 7 days earlier with four times less cases/km^2^ in regions where the lockdown was applied within 10 days of the start of the epidemic. Cumulative caseloads, cases/km^2^ and/or the number of days into an epidemic can be used to plan future localised suppression measures as part of a national post-lockdown policy. There were 350 (95% confidence interval (CI) 203) cumulative cases and 2.4 (CI 1.1) cases/km^2^ on day 8 of the regional epidemics.

## Introduction

COVID-19, defined as the respiratory disease caused by SARS-CoV-2 a novel coronavirus isolated from patients in Wuhan, was first reported on 31^st^ December 2019^1^. By early January 2020, the outbreak had spread to many cities in China, and cases began to appear outside China. On 30^th^ January 2020, the World Health Organization (WHO) declared the COVID-19 outbreak as the sixth public health emergency of international concern. By early February 2020, cases had been reported in 25 countries and in early March WHO reported that the number of cases outside China had increased 13-fold and the number of countries with cases increased threefold within a fortnight. On 11^th^ March 2020 WHO declared the novel coronavirus outbreak a global pandemic^2,3^.

The rapid spread of the disease worldwide and within some countries quickly outstripped the ability of track and trace measures to contain the COVID-19 pandemic with the currently available tests and testing facilities^4,5^. Efforts to mitigate further spread of the virus led to implementation of extraordinary public health measures in many countries, including voluntary and mandated quarantine, stopping mass gatherings, closure of educational institutes or places of work, social distancing and isolation of households, towns and cities. Eventually, complete lockdowns within national borders were enforced in an effort to suppress further spread of the virus and prevent the collapse of healthcare services, some of which had been considered centres of excellence^6,7,8,9^.

Throughout the COVID-19 pandemic, model-based predictions have played a considerable role in shaping policy makers’ decisions and recommendations on when, how and why governments have made interventions^10^. Although mathematical models can be valuable, their predictive powers are only valid within the limits of the parameters examined. In the early stages of a new pandemic, reliable parameter estimates are seldom available, often leading to the use of best guess estimates and predictions need to be constantly reviewed, as new real-world data emerge^5,8^. It cannot be guaranteed that all policy decisions, though based on current best scientific advice, are necessarily the correct decisions due to the vagaries of a rapidly changing disease picture. Indeed, the lack of clear results in countries after suppression strategies had been applied have led some to question if those measures had any value in controlling the disease^11^. One area of importance has been the timing of application of measures during the course of the pandemic. Optimal timings may differ with different disease prevention strategies. Strategies based purely on mitigation do not interrupt transmission completely but reduce the burden on health services and allow immunity to build up so that transmission eventually begins to fall. Mitigation interventions such as case isolation, home quarantine and social distancing of most at risk, applied in a three-month window around the peak of the epidemic would reduce incidence. The introduction and removal of mitigation measures needs to be timed so that transmission does not return^9^. A suppression strategy is designed to lower incidence or eliminate human-to-human transmission with interventions such as case isolation, home quarantine and more generalised social distancing, including banning mass gatherings, closure of schools and universities, up to a total lockdown of movement by all but essential workers. Such measures need to be instituted well before health services are overwhelmed^9, 12^.

As the pandemic progressed, many countries have moved their non-pharmaceutical disease prevention (NPI) strategies from mitigation measures to suppression measures or a combination of both and applied these at varying times making it difficult to determine the effects of individual measures. Ultimately, lockdown, as the last of the NPI measures, can be a point of reference for analysis of the combined effect. Although the worldwide pandemic is still active, many countries have passed the initial peak of daily new clinical cases and there is now an opportunity to analyse real data in the early stages of the pandemic in some countries. The effects of NPIs on the epidemiology of the epidemic as it first appeared in Wuhan have been examined^12-18^ and the effects of a localised lockdown in a village in Italy on disease prevalence has been reported^19^. However, as yet, there is little examination of the timing and use of national NPI strategies elsewhere. With pressure on countries to lift or ease lockdown measures and with modelling predicting the return of the pandemic after measures are eased^9^, it would be useful to know the value of measures that have been put in place, and how they might best be used in the future, should they be required again^11^.

Although, at the time of writing, Italy is still dealing with the outbreak of COVID-19 in all regions, the peak of new cases appears to have passed. On 8^th^ March 2020 a partial lockdown was imposed in the northern half of Italy on top of previous mitigating measures in the face of a flourishing epidemic. Within 2 days this lockdown was extended to the full country^20^. This paper uses actual epidemic clinical case numbers from the upward side of the epidemic curve in the various regions of Italy to analyse the effects of lockdown in each region using the national lockdown announced on 10^th^ March 2020 as a cut-off point.

## Methods

Daily data on the cumulative clinical cases and mortalities of the COVID-19 outbreak for each of the different regions of Italy were collected from the Italian Ministry of Health, COVID-19 Situation^21^.

Daily incidence rates were calculated from the data and epidemic curves constructed for each region with day one of each regional epidemic designated as the day when the incidence of clinical cases exceeds 10 cases for the first time. Epidemic peaks were described for each region and cumulative caseloads were noted on the day of peak incidence. These were standardised by dividing the cumulative caseload by regional population density, obtained from Google Search, to obtain a figure of cases/km^2^ in each region.

A national lockdown was enforced in all regions of Italy from the 10^th^ March 2020. Epidemics, as defined in this paper, had progressed in some regions by this date and in other regions, epidemics had yet to begin. The regions were divided into two groups based on when the national lockdown was enforced in relation to the start of the regional epidemic. One group contained all regions where the epidemic had not progressed beyond 10 days at the time of lockdown and the other group contained the regions where the epidemic had progressed beyond 10 days at the time of lockdown.

The two groups of regions were tested for differences in the number of cases/km^2^ and the day of the epidemic peak, using a Student T test with significance measured at p < 0.05.

Cumulative caseloads were noted on the 8^th^ day of each regions’ epidemic. These were standardised by dividing the cumulative caseload by regional population density, obtained from Google Search, to obtain a figure of cases/km^2^ in each region at day 8 of the epidemic. Mean and 95% confidence intervals were calculated from this data.

## Results

Table 1 shows that regions where lockdown was applied within 10 days of the epidemic starting had 4 times less clinical cases and peaked on average 7 days sooner than regions where lockdown was applied greater than 10 days into the epidemic and these findings were significantly different.

**Table 1.**
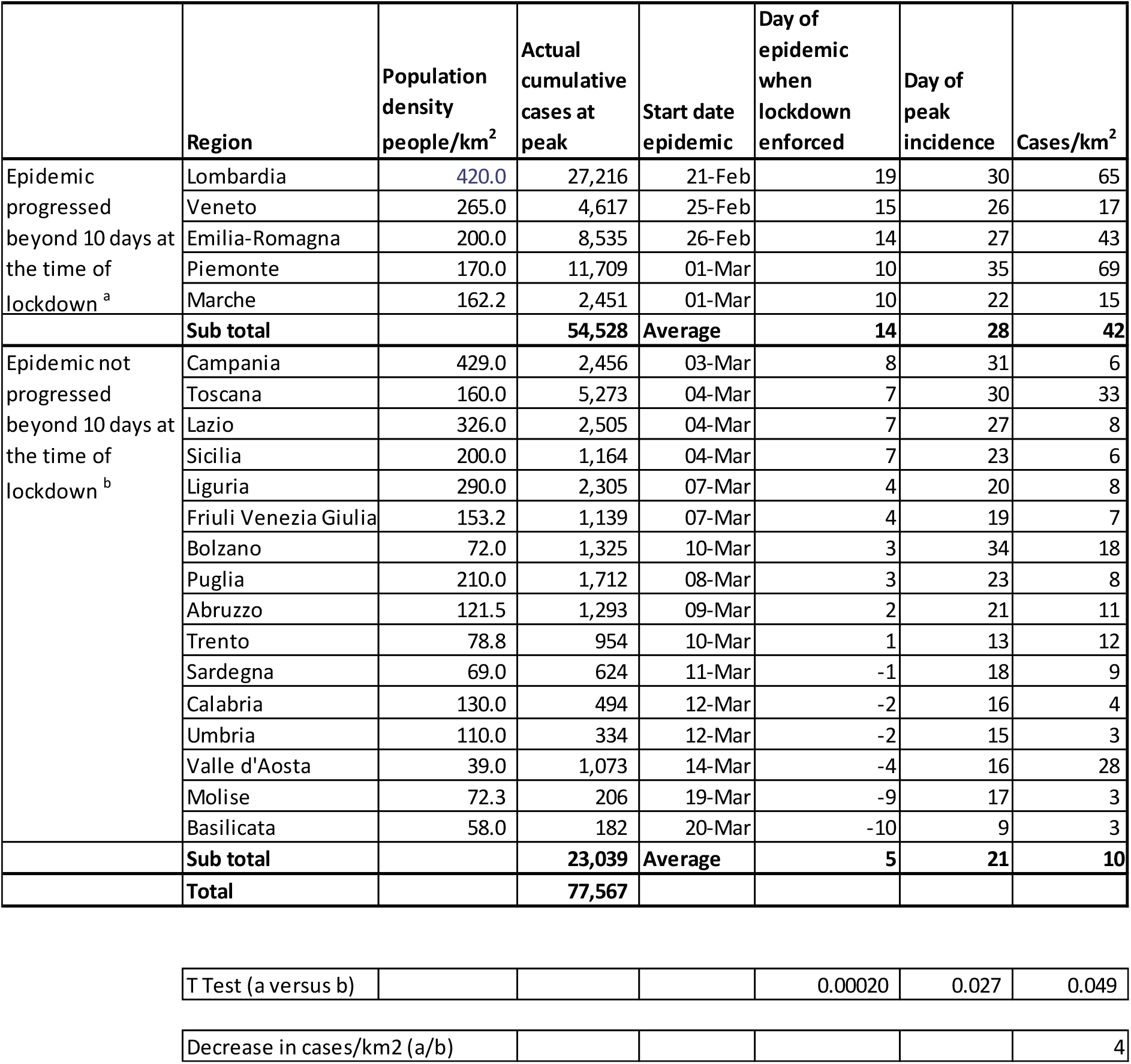
Effects of timing of lockdown on COVID-19 in the regions of Italy. Population density, Actual cumulative cases of COVID-19 at peak, Epidemic start date, Day of epidemic when lockdown was enforced, Day of peak incidence and Cases/km^2^ in the regions of Italy. (T Test a versus b P > 0.05)

Table 2 shows the actual COVID-19 incidence in the regions of Italy when the epidemic had reached day 8. On day 8 of the regional epidemics, the mean cumulative caseload was 350 (CI 203). The mean number of cases/km^2^ was 2.4 (CI 1.1).

**Table 2.** Day 8 Cumulative cases and Cases/km^2^ in the regions of Italy. Actual COVID-19 incidence in the regions of Italy when the epidemic had reached day 8. The mean cumulative caseload was 350 (CI 203). The mean number of cases/km^2^ was 2.4 (CI 1.1).

| Region | Day 8 cumulative cases | Day 8 Cases/km<sup>2</sup> |
| --- | --- | --- |
| Lombardia | 2,232 | 5 |
| Veneto | 307 | 1 |
| Emilia-Romagna | 544 | 3 |
| Piemonte | 350 | 2 |
| Marche | 302 | 2 |
| Campania | 127 | 0 |
| Toscana | 320 | 2 |
| Lazio | 150 | 0 |
| Sicilia | 83 | 0 |
| Liguria | 345 | 1 |
| Friuli Venezia Giulia | 301 | 2 |
| Bolzano | 291 | 4 |
| Puglia | 230 | 1 |
| Abruzzo | 176 | 1 |
| Trento | 385 | 5 |
| Sardegna | 134 | 2 |
| Calabria | 169 | 1 |
| Umbria | 334 | 3 |
| Valle d'Aosta | 313 | 8 |
| Molise | 103 | 1 |
| Basilicata | 151 | 3 |
| **Mean** | **350** | **2.4** |
| **Confidence Level(95.0%)** | **203** | **1.1** |

Figure 1 illustrates how earlier application of lockdown measures in a regional epidemic affected the caseload at peak incidence.

**Figure 1.**
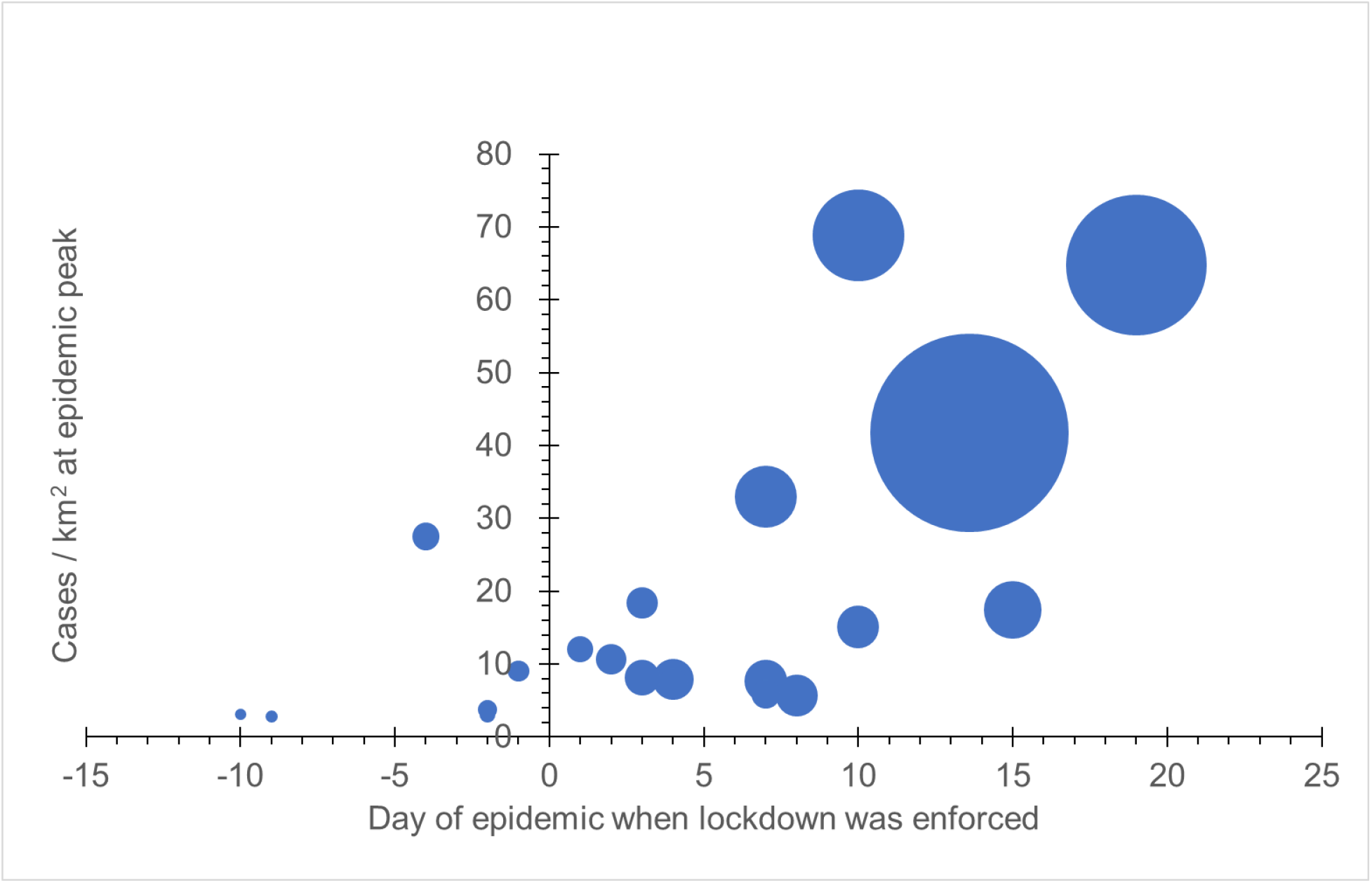
The effects of timing of lockdown on the caseload at epidemic peak. The relationship between number of cases/km^2^ at peak and the day of epidemic when lockdown was enforced for regions in Italy. The size of bubbles represents the number of cases in each region.

## Discussion

The latter half of the pandemic in Italy was not finished and lockdown measures were still in place at the time of writing this paper. Since most systematic surveillance, at least in the early phase of the pandemic has, in most countries, occurred in the hospital context, the typical delay from infection to hospitalisation means there is a 2 to 3 week lag between interventions being introduced and the impact being reflected in the reported caseload ^9^. Thus, any benefits in regions where lockdown was applied greater than 10 days into the epidemic were likely to be minimal before the peak but could have extended beyond the peak into the latter half of the epidemic. However, it is clear from these results that early lockdown and the attendant mitigating factors that preceded lockdown resulted in a much lesser caseload of clinical cases and a shortening of the time to reach epidemic peak. Studies using modelling predictions for comparison with actual case numbers found that if NPIs had been conducted one week later, then the number of cases could have shown a 3-fold increase across China^22^. This compares with the 4-fold decrease in actual case numbers following early application of lockdown in the regions of Italy in this study.

This study also follows the prediction of modelling of the more stringent preventative efforts of suppression. Modelling of suppression measures also highlight the probability of a resurgence of cases when lockdown measures are lifted^8, 9^. Measures are currently being put in place in many countries to deal with lockdown removal and include containment and suppression of any new resurgence. Previous work has shown that depending on testing capacity, the window for successful containment measures can close when the number of initial cases increases to 40^23^. Germany has already started easing lockdown restrictions and in a second wave of measures Chancellor Merkel has announced an emergency instrument or “brake” to reimpose restrictions when more than 50 people in 100,000 have been infected in the past seven days^24,25^.

This study illustrates that the window for optimal application of lockdown measures is within the first 10 days of the epidemic or when initial cases reach 350 or 2.4 cases/km^2^. Depending on the methodology adopted to define lockdown areas, these figures can be adopted to guide localised reapplication of suppression measures to deal with resurgent cases, if track and trace measures fail to control local transmission.

## Data Availability

The data used in the study is freely available on the worldwide web.

http://www.salute.gov.it/portale/nuovocoronavirus/dettaglioContenutiNuovoCoronavirus.jsp?lingua=italiano&id=5351&area=nuovoCoronavirus&menu=vuoto

## Conflict of Interest

*The authors declare that the research was conducted in the absence of any commercial or financial relationships that could be construed as a potential conflict of interest*.

## Author Contributions

All authors listed have made a substantial, direct and intellectual contribution to the work, and approved it for publication.

## Funding

The authors declare that the research was conducted in the absence of any commercial or financial relationships that could be construed as a potential conflict of interest.

## Acknowledgments

Thanks to Dr Clare Kettle for help with tables and figures.

## Notes

### Competing Interest Statement

The authors have declared no competing interest.

